# The Impact of Loneliness on Depression, Mental Health Days, and Physical Health

**DOI:** 10.1101/2025.02.02.25321551

**Authors:** Oluwasegun Akinyemi, Waliah Abdulrazaq, Mojisola Fasokun, Fadeke Ogunyankin, Seun Ikugbayigbe, Uzoamaka Nwosu, Mariam Michael, Kakra Hughes, Temitope Ogundare

## Abstract

**Introduction:** Loneliness is a significant public health concern and a well-established social determinant of health, affecting both mental and physical well-being. It has been linked to an increased risk of depression, anxiety, cardiovascular disease, and premature mortality. Despite growing awareness, loneliness remains an underrecognized and undertreated factor influencing population health.

**Objective:** This study examines the impact of loneliness on the likelihood of being diagnosed with depression, as well as its association with self-reported poor mental and physical health days.

**Methods:** Data were analyzed from the Behavioral Risk Factor Surveillance System (BRFSS) (2016–2023). The primary exposure was self-reported loneliness, captured through the question, “How often do you feel lonely?” with responses ranging from “Always” to “Never.” Main outcomes included depression diagnosis, poor mental health days, and poor physical health days. Covariates included age, race, gender, marital status, education, employment, state, year, metropolitan status, and language spoken at home. Inverse Probability Weighting (IPW) was used to estimate the Average Treatment Effect (ATE), accounting for confounders and state and year fixed effects. Sampling weights ensured national representativeness, and robust standard errors accounted for clustering by state.

**Results:** Among 47,026 participants, 82.4% reported experiencing some degree of loneliness, with 6.2% feeling “Always” lonely, 8.3% feeling “Usually” lonely, 37.9% feeling “Sometimes” lonely, and 29.9% feeling “Rarely” lonely. In contrast, 17.7% of participants reported “Never” feeling lonely. For further analysis, 2,609 individuals who reported feeling lonely were matched with 2,609 individuals who reported “Never” feeling lonely”, forming a balanced comparison group. The “Always Lonely” population was predominantly White (64.5%) and female (55.0%), with the majority aged 45–64 years. Loneliness was significantly associated with an increased likelihood of depression diagnosis, with a 39.3% percentage-point increase for those reporting Always lonely (ATE = 0.39, 95% CI: 0.34–0.44, p < 0.001). Loneliness was also associated with a 10.9-day increase in poor mental health days (ATE = 10.9, 95% CI: 9.8–11.9, p < 0.001) and a 5.0 day increase in poor physical health days (ATE = 5.0, 95% CI: 3.8–6.1, p < 0.001).

**Conclusions:** Loneliness is a strong predictor of depression and poor mental and physical health. Interventions addressing social isolation could mitigate the negative health impacts associated with loneliness, improving population health outcomes.

## Introduction

Loneliness, a growing public health challenge in the United States, has profound implications for depression, mental health, and physical well-being(1–3). This silent epidemic affects approximately 36% of Americans, who report feeling lonely “frequently” or “all the time,” with higher rates among young adults (61%) and mothers with young children (51%)(4). The prevalence varies by sex, with women reporting loneliness at higher rates than men(4). Loneliness has been linked to a 26% increase in the risk of premature death, comparable to the impact of obesity and smoking(1, 4). These health implications underscore the importance of addressing loneliness as a critical public health issue.

Loneliness has been strongly linked to an increased risk of depression, with studies showing a bidirectional relationship between the two(5, 6). Social isolation amplifies feelings of hopelessness, sadness, and lack of purpose, contributing to the development and exacerbation of depressive disorders(7). The prevalence of depression among lonely individuals is estimated to be 15-30% higher compared to those with strong social connections(8). Furthermore, mental health days, defined as days when an individual’s mental or emotional health interferes with daily activities, are significantly affected by loneliness. Research indicates that lonely individuals report an average of two to three more poor mental health days per month compared to their socially connected counterparts(7, 9). These days are characterized by increased stress, anxiety, and emotional instability, impacting the overall quality of life and productivity(10).

Physical health days, which refer to days when physical health issues hinder regular activities, are also influenced by loneliness. Lonely individuals report an average of one to two more poor physical health days per month, often experiencing symptoms related to cardiovascular issues, weakened immune function, and chronic inflammation(4, 10, 11). Studies have shown that lonely individuals are at higher risk of developing chronic physical conditions(4, 10). The cumulative burden of loneliness on health outcomes, particularly its significant impact on depression, mental health days, and physical health days, underscores its importance as a critical public health priority.

Despite various initiatives to address loneliness, such as the Campaign to End Loneliness and local community-based interventions that aim to foster social connections through support groups, mentorship programs, and digital platforms, significant gaps remain in our understanding of its specific impacts on depression, mental health days, and physical health days(12–14). Current interventions often fail to account for the complex interplay between these health outcomes and loneliness, particularly across diverse demographic groups(9, 14). Additionally, the prevalence and impact of loneliness are closely tied to social determinants of health(15). Factors such as socioeconomic status, educational attainment, and access to social support networks play pivotal roles in shaping the experience of loneliness and its subsequent health outcomes(15).

This study aims to quantify the impact of loneliness on depression, mental health days, and physical health days in the United States. The primary objective is to examine the association between loneliness and health outcomes, adjusting for key social determinants of health. A secondary objective is to evaluate differences in the burden of loneliness across demographic subgroups, including sex, age, and race/ethnicity. We hypothesize that higher levels of loneliness are significantly associated with an increased likelihood of depression, more poor mental and physical health days, and poorer overall health status. By addressing these gaps, this study seeks to provide actionable insights to inform public health strategies aimed at mitigating the impact of loneliness on health outcomes.

## Methodology

### Study Design and Data Source

This study utilized data from the Behavioral Risk Factor Surveillance System (BRFSS) spanning January 2016 to December 2023(16). The BRFSS is a cross-sectional, nationally representative survey conducted annually to collect data on health-related risk behaviors, chronic health conditions, and preventive health practices among U.S. adults. A total of 321,106 respondents were included in the analysis to ensure a robust sample for generalizability.

### Primary Exposure Variable

The primary exposure variable was self-reported loneliness, measured through the question: *“How often do you feel lonely?”* Respondents were provided with five response options: Always, Usually, Sometimes, Rarely, and Never. The variable was categorized as ordinal for analysis, with “Always” and “Usually” indicating frequent loneliness, “Sometimes” as moderate loneliness, and “Rarely” and “Never” as low or no loneliness. Responses of “Don’t know/Not sure” or “Refused” were excluded.

### Outcome Variables

1. **Depression**: Self-reported depression was assessed using the question: *“(Ever told) (you had) a depressive disorder (including depression, major depression, dysthymia, or minor depression)?”* Responses were coded as a binary variable, where “Yes” indicated a diagnosis of depression and “No” indicated no diagnosis. “Don’t know/Not sure” and “Refused” responses were excluded.
2. **Mental Health Days**: The number of poor mental health days was derived from responses to the question: *“For how many days during the past 30 days was your mental health not good?”* Responses ranged from 0 to 30 days and were treated as a continuous variable. Non-responses and “Don’t know/Not sure” or “Refused” responses were excluded.
3. **Physical Health Days**: Physical health days were assessed using the question: *“For how many days during the past 30 days was your physical health not good?”* Responses ranged from 0 to 30 days and were treated as a continuous variable, with exclusions applied to missing or unclear responses.

### Covariates and Confounder Adjustment

The analysis adjusted for a comprehensive set of demographic and socioeconomic covariates, including race/ethnicity, gender, age, education level, marital status, employment status, state, year, metropolitan status, and language spoken at home. These covariates were included to control for potential confounding effects on both the exposure and outcomes.

### Statistical Analysis

Inverse Probability Weighting (IPW) was employed to estimate the Average Treatment Effect (ATE) of loneliness on depression, mental health days, and physical health days. This approach minimizes selection bias by creating a pseudo-population with balanced covariates. Propensity scores were calculated using logistic regression, incorporating all covariates. Sampling weights provided by BRFSS were applied to ensure national representativeness. State and year fixed effects were included to account for regional and temporal heterogeneity.

### Robustness and Precision

Robust standard errors clustered at the state level were used to account for within-state correlations and ensure accurate confidence intervals. The model output included ATE estimates, potential outcome means (POmeans), and 95% confidence intervals, with statistical significance determined at p < 0.05.

### Ethics and Data Access

The study was conducted in accordance with the ethical standards of the institutional and/or national research committee and with the 1964 Helsinki declaration and its later amendments or comparable ethical standards. Institutional Review Board approval was waived because the study was carried out on a national database that contained de-identified data and did not require informed consent or direct participation of patients.

## Results: Baseline Characteristics

Table 1 presents the baseline characteristics of individuals reporting loneliness (“Always Lonely”) compared to those who reported “Never Lonely” in both unmatched and matched samples. The unmatched sample comprised 11,206 participants, while the matched cohort included 5,218 participants evenly distributed between the two groups. Key differences were observed in demographic, socioeconomic, and health-related variables.

**Table 1:** Baseline Covariate Balance for Unmatched and Matched Samples.

|  | Unmatched Sample (N=11,206) |  | Matched Sample (n=5,218) |  |  |
| --- | --- | --- | --- | --- | --- |
| Variable | Always Lonely<br>(n=2,907) | Never Lonely<br>(n=8,299) | Always Lonely<br>(n=2,609) | Never Lonely<br>(n=2,609) | % Bias<br>Reduction |
| **Age (Yr.)** |  |  |  |  |  |
| 18-44Yr. | 963.96 (33.16%) | 2352.77 (28.35%) | 861.75 (33.03%) | 973.94 (37.33%) | 0.104 |
| 45-64Yr. | 1223.85 (42.1%) | 3145.32 (37.9%) | 1097.87 (42.08%) | 928.02 (35.57%) | -54.8 |
| >64Yr. | 718.9 (24.73%) | 2800.91 (33.75%) | 649.38 (24.89%) | 707.04 (27.1%) | 75.5 |
| **Race/Ethnicity** |  |  |  |  |  |
| White | 1866.0 (64.19%) | 5996.03 (72.25%) | 1682.02 (64.47%) | 1767.6 (67.75%) | 59.4 |
| Black | 315.12 (10.84%) | 781.77 (9.42%) | 283.34 (10.86%) | 266.12 (10.2%) | 53.3 |
| Hispanic | 314.25 (10.81%) | 664.75 (8.01%) | 282.29 (10.82%) | 229.59 (8.8%) | 27.7 |
| Other | 411.63 (14.16%) | 855.63 (10.31%) | 361.09 (13.84%) | 345.95 (13.26%) | 84.7 |
| Female (ref. Male) | 1590.13 (54.7%) | 4864.87 (58.62%) | 1435.99 (55.04%) | 1638.71 (62.81%) | -97.9 |
| **Marital Status** |  |  |  |  |  |
| Married | 543.9 (18.71%) | 4801.8 (57.86%) | 494.67 (18.96%) | 646.51 (24.78%) | 85.1 |
| Divorced | 732.85 (25.21%) | 1100.45 (13.26%) | 667.64 (25.59%) | 550.5 (21.1%) | 62.4 |
| Widowed | 461.92 (15.89%) | 624.08 (7.52%) | 411.96 (15.79%) | 375.7 (14.4%) | 83.3 |
| Separated | 163.08 (5.61%) | 167.64 (2.02%) | 146.89 (5.63%) | 102.79 (3.94%) | 52.7 |
| Never Married | 872.97 (30.03%) | 1195.06 (14.4%) | 770.44 (29.53%) | 756.87 (29.01%) | 96.7 |
| Member of an unmarried couple | 131.98 (4.54%) | 409.97 (4.94%) | 117.14 (4.49%) | 176.89 (6.78%) | -471.3 |
| Employed (ref. Unemployed) | 847.1 (29.14%) | 3916.3 (47.19%) | 767.57 (29.42%) | 1007.6 (38.62%) | 49 |
| **Education Level** |  |  |  |  |  |
| High School or Less | 1478.21 (50.85%) | 2790.12 (33.62%) | 1320.94 (50.63%) | 1000.03 (38.33%) | 28.6 |
| College | 896.23 (30.83%) | 2429.12 (29.27%) | 805.92 (30.89%) | 845.32 (32.4%) | 3.3 |
| Advanced | 532.56 (18.32%) | 3079.76 (37.11%) | 482.14 (18.48%) | 763.65 (29.27%) | 42.6 |
| **Insurance** |  |  |  |  |  |
| Private | 581.69 (20.01%) | 3527.07 (42.5%) | 520.76 (19.96%) | 925.15 (35.46%) | 31.1 |
| Medicare | 936.64 (32.22%) | 2648.21 (31.91%) | 842.45 (32.29%) | 810.88 (31.08%) | -283.3 |
| Medicaid | 682.85 (23.49%) | 793.38 (9.56%) | 611.03 (23.42%) | 400.48 (15.35%) | 42.1 |
| Self-Pay | 262.21 (9.02%) | 390.05 (4.7%) | 239.25 (9.17%) | 115.32 (4.42%) | -10 |
| Other | 443.32 (15.25%) | 941.11 (11.34%) | 395.79 (15.17%) | 357.43 (13.7%) | 62.4 |
| Spanish Speaking (ref. English speaking) | 206.98 (7.12%) | 361.84 (4.36%) | 189.15 (7.25%) | 118.19 (4.53%) | 1.2 |
| Metro (ref. non-Metro) | 915.12 (31.48%) | 2456.5 (29.6%) | 838.01 (32.12%) | 815.57 (31.26%) | 54.2 |
| Urban (ref. Rural) | 492.16 (16.93%) | 1235.72 (14.89%) | 446.92 (17.13%) | 402.83 (15.44%) | 17.3 |

In the unmatched sample, individuals in the “Always Lonely” group were younger, with 33.2% aged 18–44 years, compared to 28.4% in the “Never Lonely” group. After matching, the age distribution was balanced, with 33.0% of the “Always Lonely” group and 37.3% of the “Never Lonely” group aged 18–44 years, achieving significant bias reduction. Similarly, differences in race/ethnicity were apparent pre-matching, with 64.2% of the “Always Lonely” group identifying as White compared to 72.3% of the “Never Lonely” group. Matching reduced this imbalance, yielding 64.5% and 67.8% White participants in the respective groups.

Marital status also displayed substantial differences. In the unmatched sample, the “Always Lonely” group had higher proportions of divorced (25.2%) and widowed (15.9%) individuals compared to the “Never Lonely” group (13.3% divorced and 7.5% widowed). Post-matching, these differences narrowed, with 25.6% divorced and 15.8% widowed in the “Always Lonely” group, compared to 21.1% divorced and 14.4% widowed in the “Never Lonely” group.

Socioeconomic disparities were evident in education and employment. In the unmatched sample, 50.9% of the “Always Lonely” group had a high school education or less, compared to 33.6% in the “Never Lonely” group. Post-matching, this gap narrowed to 50.6% and 38.3%, respectively. Employment status differences also improved, with 29.4% employed in the “Always Lonely” group and 38.6% in the “Never Lonely” group post-matching, compared to a wider gap pre-matching (29.1% vs. 47.2%).

Bias reduction across all covariates confirms the effectiveness of the matching process, ensuring comparability between the groups for subsequent analysis (Table 1).

### Average Treatment Effects of Loneliness on Depression Incidence

Table 2 outlines the estimated ATE of loneliness on the likelihood of being diagnosed with depression across varying levels of self-reported loneliness. The analysis highlights a clear dose-response relationship, where higher levels of loneliness are associated with a progressively greater risk of depression.

**Table 2:**
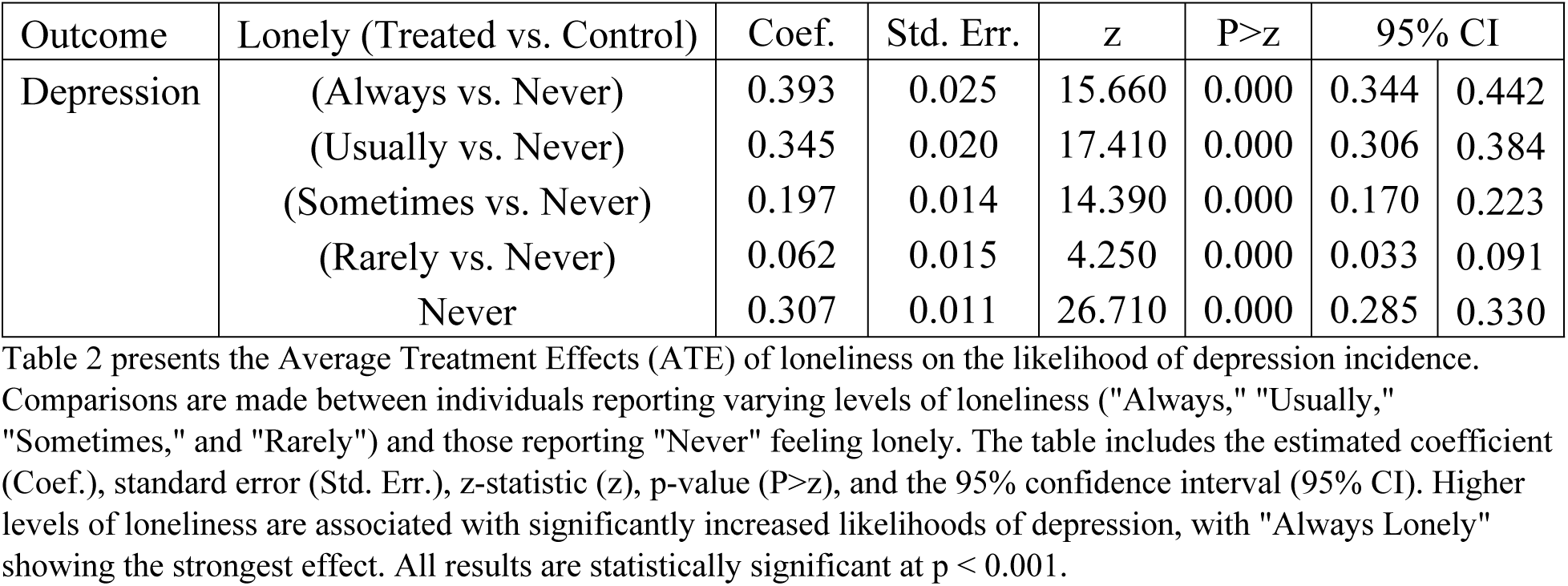
Average Treatment Effects of Loneliness on Depression Incidence.

Individuals who reported feeling “Always Lonely” experienced a significant 39.3 percentage-point increase in the likelihood of depression compared to those who reported “Never Lonely” (Coef. = 0.393, Std. Err. = 0.025, z = 15.660, p < 0.001, 95% CI: 0.344–0.442). Those who reported “Usually Lonely” also showed a substantial increase in depression risk, with an estimated 34.5 percentage-point rise (Coef. = 0.345, Std. Err. = 0.020, z = 17.410, p < 0.001, 95% CI: 0.306–0.384). Similarly, participants in the “Sometimes Lonely” category had a 19.7 percentage-point increase in depression likelihood compared to “Never Lonely” individuals (Coef. = 0.197, Std. Err. = 0.014, z = 14.390, p < 0.001, 95% CI: 0.170–0.223).

Even individuals who reported feeling “Rarely Lonely” demonstrated an elevated risk of depression, with a 6.2 percentage-point increase relative to the “Never Lonely” group (Coef. = 0.062, Std. Err. = 0.015, z = 4.250, p < 0.001, 95% CI: 0.033–0.091). The “Never Lonely” group exhibited a baseline depression incidence of 30.7% (Coef. = 0.307, Std. Err. = 0.011, z = 26.710, p < 0.001, 95% CI: 0.285–0.330).

These findings underscore the graded association between loneliness and depression, with higher loneliness levels correlating with significantly increased risks of depression (Table 2).

### Average Treatment Effects of Loneliness on Self-Reported Poor Mental Health Days per Month

Table 3 presents the ATE of loneliness on self-reported poor mental health days per month, demonstrating a strong association between higher levels of loneliness and increased mental health burden. The analysis reveals a dose-response relationship, with greater loneliness corresponding to significantly more poor mental health days.

**Table 3:**
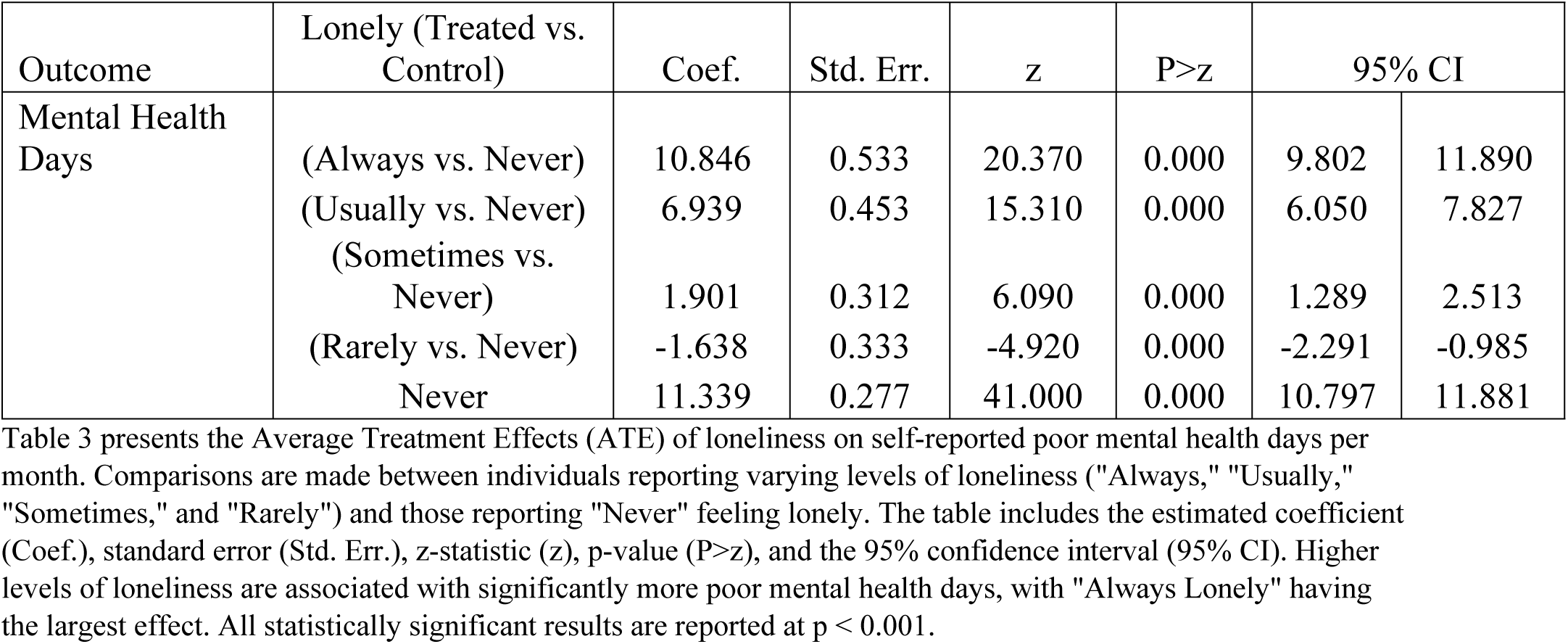
Average Treatment Effects of Loneliness on Self-Reported Poor Mental Health Days per Month.

Individuals reporting “Always Lonely” experienced an average increase of 10.85 poor mental health days per month compared to those who reported “Never Lonely” (Coef. = 10.846, Std. Err. = 0.533, z = 20.370, p < 0.001, 95% CI: 9.802–11.890). Similarly, those in the “Usually Lonely” category reported 6.94 additional poor mental health days per month relative to the “Never Lonely” group (Coef. = 6.939, Std. Err. = 0.453, z = 15.310, p < 0.001, 95% CI: 6.050–7.827).

Participants who identified as “Sometimes Lonely” reported a smaller but still significant increase of 1.90 poor mental health days per month compared to the “Never Lonely” group (Coef. = 1.901, Std. Err. = 0.312, z = 6.090, p < 0.001, 95% CI: 1.289–2.513). Interestingly, individuals who reported feeling “Rarely Lonely” experienced a slight reduction in poor mental health days, with an average decrease of 1.64 days per month relative to the “Never Lonely” group (Coef. = −1.638, Std. Err. = 0.333, z = −4.920, p < 0.001, 95% CI: −2.291 to −0.985).

The “Never Lonely” group reported an average of 11.34 poor mental health days per month (Coef. = 11.339, Std. Err. = 0.277, z = 41.000, p < 0.001, 95% CI: 10.797–11.881).

These findings demonstrate the significant mental health burden associated with loneliness, with the most pronounced effects observed in individuals reporting persistent loneliness (Table 3).

### Average Treatment Effects of Loneliness on Self-Reported Poor Physical Health Days per Month

Table 4 examines the ATE of loneliness on self-reported poor physical health days per month, revealing a significant association between higher levels of loneliness and increased physical health burden. The results demonstrate a dose-response relationship, with individuals reporting more loneliness experiencing an increase in more poor physical health days.

**Table 4:**
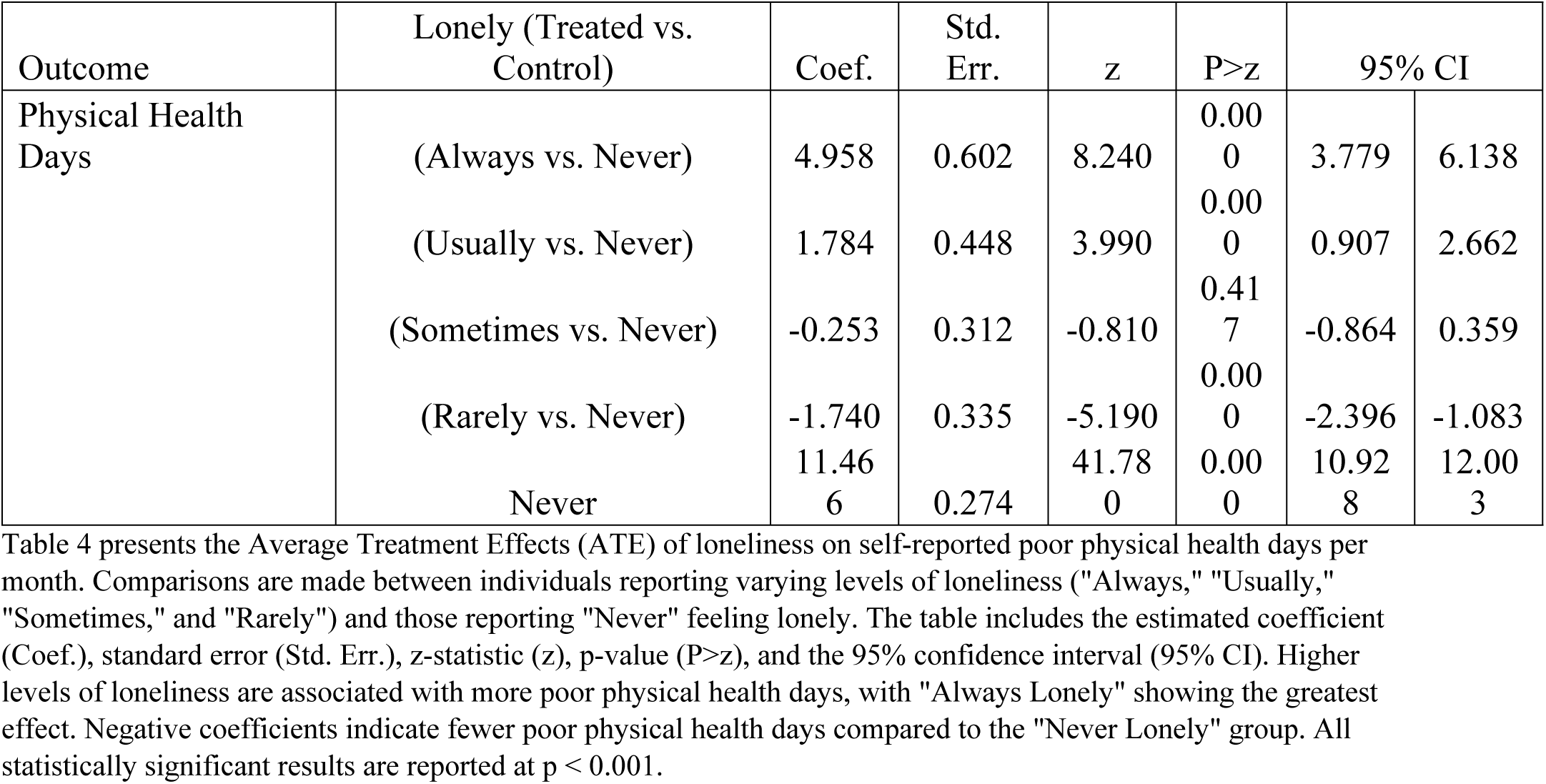
Average Treatment Effects of Loneliness on Self-Reported Poor Physical Health Days per Month.

Participants who reported being “Always Lonely” experienced an average increase of 4.96 poor physical health days per month compared to those who reported “Never Lonely” (Coef. = 4.958, Std. Err. = 0.602, z = 8.240, p < 0.001, 95% CI: 3.779–6.138). Individuals in the “Usually Lonely” group reported 1.78 additional poor physical health days per month compared to the “Never Lonely” group (Coef. = 1.784, Std. Err. = 0.448, z = 3.990, p < 0.001, 95% CI: 0.907–2.662).

In contrast, individuals reporting “Sometimes Lonely” showed no significant difference in poor physical health days compared to the “Never Lonely” group (Coef. = −0.253, Std. Err. = 0.312, z = −0.810, p = 0.417, 95% CI: −0.864–0.359). Interestingly, participants who identified as “Rarely Lonely” reported significantly fewer poor physical health days, with an average reduction of 1.74 days compared to the “Never Lonely” group (Coef. = −1.740, Std. Err. = 0.335, z = −5.190, p < 0.001, 95% CI: −2.396 to −1.083).

The “Never Lonely” group reported an average of 11.47 poor physical health days per month (Coef. = 11.466, Std. Err. = 0.274, z = 41.780, p < 0.001, 95% CI: 10.928–12.003).

These findings highlight the physical health burden associated with persistent loneliness, with the strongest effects observed in those experiencing chronic loneliness (Table 4).

## Discussion

Our study highlights the profound impact of loneliness on mental and physical health outcomes, revealing significant associations with increased rates of depression diagnoses, poor mental health days, and poor physical health days. These findings underscore the importance of addressing loneliness as a critical public health concern.

### Loneliness and Depression

We observed a graded association between loneliness and depression, with individuals who reported “Always” feeling lonely experiencing a 39.3 percentage-point increase in the likelihood of depression diagnosis compared to those who reported “Never” feeling lonely. This significant increase aligns with existing literature that links social isolation and loneliness to heightened risks of depression(17, 18). Pathophysiologically, loneliness activates the hypothalamic-pituitary-adrenal (HPA) axis, leading to chronic stress and dysregulation of neurotransmitters, such as serotonin and dopamine, which are critical for mood regulation(19–21). Additionally, loneliness impairs cognitive processes and fosters feelings of hopelessness, further exacerbating depressive symptoms(19, 22).

Comparatively, studies have shown similar trends; for instance, Cacioppo et al. (2015) demonstrated that loneliness was a strong predictor of depression onset, independent of other psychosocial factors(23). Our findings not only confirm these associations but also emphasize the magnitude of the effect in a nationally representative cohort.

The societal and public health implications of this association are substantial. Depression is a leading cause of disability worldwide, contributing to increased healthcare costs, reduced productivity, and heightened suicide rates(5, 24). Addressing loneliness could therefore play a pivotal role in mitigating these broader societal burdens. Interventions such as social prescribing, peer support programs, and digital platforms fostering social connection may hold promise in reducing loneliness and its depressive consequences(25–27).

### Loneliness and Poor Mental Health Days

Individuals experiencing loneliness also reported a marked increase in the number of poor mental health days. Those who “Always” felt lonely had an average increase of 10.9 poor mental health days per month compared to non-lonely individuals. This finding aligns with previous research linking loneliness to chronic psychological distress, heightened anxiety, and emotional dysregulation(10, 28, 29). The dose-response relationship observed in our study reinforces the notion that the severity of loneliness directly correlates with its impact on mental health.

Mechanistically, loneliness induces chronic stress, which disrupts the balance of the autonomic nervous system and promotes systemic inflammation(30–32). These physiological changes exacerbate feelings of distress and reduce resilience to everyday stressors(31, 32). Moreover, loneliness often co-occurs with poor sleep quality, which further impairs mental well-being(33–36).

From a public health perspective, the increase in poor mental health days among lonely individuals translates to reduced quality of life and increased reliance on mental health services. Policies promoting community engagement, mental health education, and stigma reduction around loneliness could mitigate its adverse effects. Schools, workplaces, and healthcare providers should also prioritize mental health screenings and interventions targeting socially isolated individuals.

### Loneliness and Poor Physical Health Days

Loneliness was also significantly associated with an increase in poor physical health days, with individuals who “Always” felt lonely reporting 5.0 additional days of poor physical health per month. This finding underscores the interconnectedness of mental and physical health. Chronic loneliness has been linked to adverse health behaviors such as poor diet, lack of exercise, and substance use, which contribute to physical health deterioration(1, 10, 37). Furthermore, prolonged activation of the HPA axis and systemic inflammation in lonely individuals accelerates the development of chronic conditions such as cardiovascular disease, diabetes, and immune dysfunction(30, 38).

Comparable studies have highlighted similar findings; Holt-Lunstad et al. (2010) demonstrated that loneliness increases mortality risk to levels comparable with smoking or obesity(39). These findings underline the importance of integrating physical health interventions with strategies addressing loneliness.

### Broader Implications

The implications of these findings extend beyond individual health to broader societal and economic consequences. Loneliness contributes to increased healthcare utilization, productivity loss, and societal costs associated with chronic disease management. Addressing loneliness could therefore yield significant economic and social benefits.

### Policy Implications and Recommendations

Tackling loneliness requires a multifaceted approach. At the policy level, national campaigns to raise awareness about the health impacts of loneliness could reduce stigma and encourage affected individuals to seek help. Healthcare providers should integrate loneliness screening into routine care, especially for high-risk groups such as the elderly, unemployed, and those with chronic illnesses. Community-level initiatives, such as promoting volunteerism, creating accessible social hubs, and implementing neighborhood-based programs, can foster social connections.

Technological solutions, such as online support groups and telehealth interventions, could also play a critical role, particularly in rural or underserved areas. Educational programs in schools and workplaces could emphasize the importance of social relationships and equip individuals with tools to build and maintain meaningful connections.

### Limitations

This study, while robust in design and methodology, has several limitations that should be acknowledged. First, although the use of IPW helps mitigate confounding, unmeasured confounders, such as personality traits, life events, or social support, may still influence the observed associations.

Second, self-reported measures for loneliness, depression, and poor mental and physical health days may introduce bias due to recall inaccuracies or social desirability. Respondents may underreport or overreport these experiences based on subjective perceptions or cultural norms, potentially impacting the accuracy of the findings.

Third, while the study adjusts for numerous demographic and socioeconomic covariates, the BRFSS lacks detailed information on structural determinants of loneliness, such as neighborhood characteristics, access to healthcare, or social capital. These factors could further contextualize the relationship between loneliness and health outcomes but remain unaccounted for in the analysis.

Fourth, the categorization of loneliness into ordinal levels (e.g., “Always,” “Usually,” “Sometimes”) may oversimplify a complex emotional experience. Loneliness is a multifaceted construct that varies across contexts and time, and this study does not capture its dynamic nature or intensity over time.

Finally, while the BRFSS is a nationally representative dataset, its reliance on telephone surveys may exclude vulnerable populations, such as individuals without access to telephones or those experiencing severe social isolation. This could lead to underrepresentation of individuals at the highest risk of loneliness and related health impacts.

Despite these limitations, the study provides valuable insights into the significant associations between loneliness and adverse health outcomes. Future research should consider longitudinal designs, more nuanced measures of loneliness, and the inclusion of structural and contextual factors to further elucidate these relationships.

## Conclusion

In conclusion, our study provides robust evidence of the detrimental effects of loneliness on depression, mental health, and physical health. By recognizing loneliness as a significant public health issue, we can implement targeted interventions to improve population health outcomes and reduce the societal burden of loneliness. Future research should explore the long-term effectiveness of such interventions and identify strategies to sustain social connections in an increasingly digital world.

## Data Availability

All relevant data are within the manuscript and its Supporting Information files.

## Notes

### Competing Interest Statement

The authors have declared no competing interest.

